# SARS-CoV-2 Antibody Response in Patients Undergoing Kidney Transplantation

**DOI:** 10.1101/2021.07.25.21261066

**Authors:** Michelle Lubetzky, Ashley Sukhu, Zhen Zhao, Sophie Rand, Vijay Sharma, Samuel Sultan, Zoe Kapur, Shady Albakry, Nataliya Hauser, Jehona Marku-Podvorica, Rebecca Craig-Schapiro, John R. Lee, Thalia Salinas, Meredith Aull, Sandip Kapur, Melissa Cushing, Darshana M. Dadhania

## Abstract

The response of the immune system to COVID-19 in end stage kidney disease patients who undergo kidney transplantation has yet to be described. We report data on 72 patients who underwent SARS-CoV-2 antibody testing both before and after kidney transplantation and were followed for a median of 186 days (range 83, 277). Of the 25 patients with a positive antibody test at the time of transplant, 17 (68%) remained positive after transplantation. Patients were significantly more likely to have a persistently positive test if they reported a symptomatic COVID-19 infection prior to transplant (p=0.01). SARS-CoV-2 IgG index values were measured in a subset of kidney transplant recipients and compared to wait -listed dialysis patients. These assays demonstrated a more significant decline in IgG (58% versus 14% p = 0.008) in transplant recipients when compared to dialysis patients tested during the same time period. Additional analysis of the quality of the immune response measuring the binding of SARS-CoV-2 antibodies to the receptor-binding domain (RBD binding), the antibody neutralizing capability, and the antibody avidity demonstrated a more pronounced effect when comparing pre-transplant values to post-induction therapy/post transplant values. The attenuated IgG response seen in transplant patients compared to dialysis patients after induction therapy requires further study. These data have important implications for post-transplant management of vaccinated dialysis patients.

## Introduction

Currently, over 33 million cases of COVID-19 have been reported in the United States. Recent population-based studies have looked specifically at the prevalence of antibodies to SARS-CoV-2 in New York and estimate that anywhere from 6% to 22% of New Yorkers have SARS-CoV-2 antibodies^1-3^. Estimates in the dialysis population have been even higher and range from 6.9%, to 69%^4,5^. With past infection rates this high, patients are presenting for kidney transplants with a prior history of COVID-19 illness, yet data are lacking on the durability of the immune response after induction immunosuppression. To better understand the changes that occur with immunosuppressive therapies, we followed SARS-CoV-2 antibody presence in transplant recipients before and after kidney transplant, specifically after the initiation of lymphocyte depleting induction immunotherapy and maintenance immunosuppression. We also compared SARS-CoV-2 antibody response in our cohort of transplant patients to a cohort of wait-listed dialysis patients over a similar time period to determine the effects of immunosuppression on antibody response.

## Materials and Methods

### Patient Cohorts, Data Collection and Analysis

This study was approved by the Weill Cornell Medicine Institutional Review Board protocol # 1207012637 entitled Utilizing a Transplant Database for Quality Assessment and Performance Improvement and Clinical Outcomes and protocol # 20-05022154 entitled Impact of COVID-19 Illness on Kidney Transplant Candidates and Recipients.

Medical records of all patients admitted for kidney transplant from May 28, 2020 until December 12, 2020 were reviewed. Inclusion criteria for the study was any patient admitted for kidney transplant who had SARS-CoV-2 antibody testing both before and after kidney transplantation (n=72). All patients were screened at the time of transplant for signs or symptoms of COVID-19 infection and tested by real time PCR (RT-PCR). Following transplantation, patients were screened for symptoms of COVID-19 illness, exposure to COVID-19 illness, and tested for SARS-CoV-2 antibodies post-transplant at one time-point post transplant. A flowsheet and description of the study cohort is shown in Figure 1. Demographic data were collected on patients who met inclusion criteria (Table 1).

**Table 1:** Characteristics of Patients Tested for SARS-CoV-2 Antibody.

| Characteristic | Pre<br>Transplant<br>Antibody<br>Positive | Pre<br>Transplant<br>Antibody<br>Negative | P Value (antibody<br>positive vs.<br>antibody negative) |
| --- | --- | --- | --- |
|  | N=25 | N=47 |  |
| Age, mean (SD) | 54.9 (13) | 55.3 (15) | 0.8 |
| Gender N, (%), Female | 10 (40) | 22 (47) | 0.65 |
| Race N (%) |  |  |  |
| Caucasian | 8 (32) | 20 (43) | 0.45 |
| Black | 9 (36) | 8 (17) | 0.09 |
| Hispanic | 3 (12) | 10 (21) | 0.52 |
| Asian | 1 (4) | 7 (15) | 0.05 |
| Type of Transplant, N (%) Living Donor | 16 (64) | 25 (53) | 0.46 |
| ATG induction | 23 (92) | 39 (83) | 0.48 |
| **Steroid Maintenance** | **1 (4)** | **11 (23)** | **0.05** |
| **Cause of ESKD, N (%)** |  |  |  |
| **HTN** | **8 (32)** | **14 (30)** | **1** |
| **DM** | **5 (20)** | **7 (15)** | **0.36** |
| **GN** | **3 (12)** | **8 (17)** | **0.52** |
| **SARS-CoV-2 PCR Positive Pre-Transplant** | **0(0)** | **0(0)** | **1** |
| **Mean (SD) 1 month creatinine (mg/dL)** | **1.7 (0.6)** | **1.6 (0.6)** | **0.26** |
| **Mean (SD) 3 month creatinine (mg/dL)** | **1.5 (0.6)** | **1.5 (0.6)** | **0.14** |
| **Mean (SD) 6 months creatinine (mg/dL)** | **1.4 (0.3)** | **1.4 (0.3)** | **0.26** |
| **Graft Loss, N (%)** | **1 (3.6)** | **0 (0)** | **0.35** |
| **COVID-19 Post Transplant, N (%)** | **0 (0)** | **3 (6)** | **0.55** |

**Figure 1.**
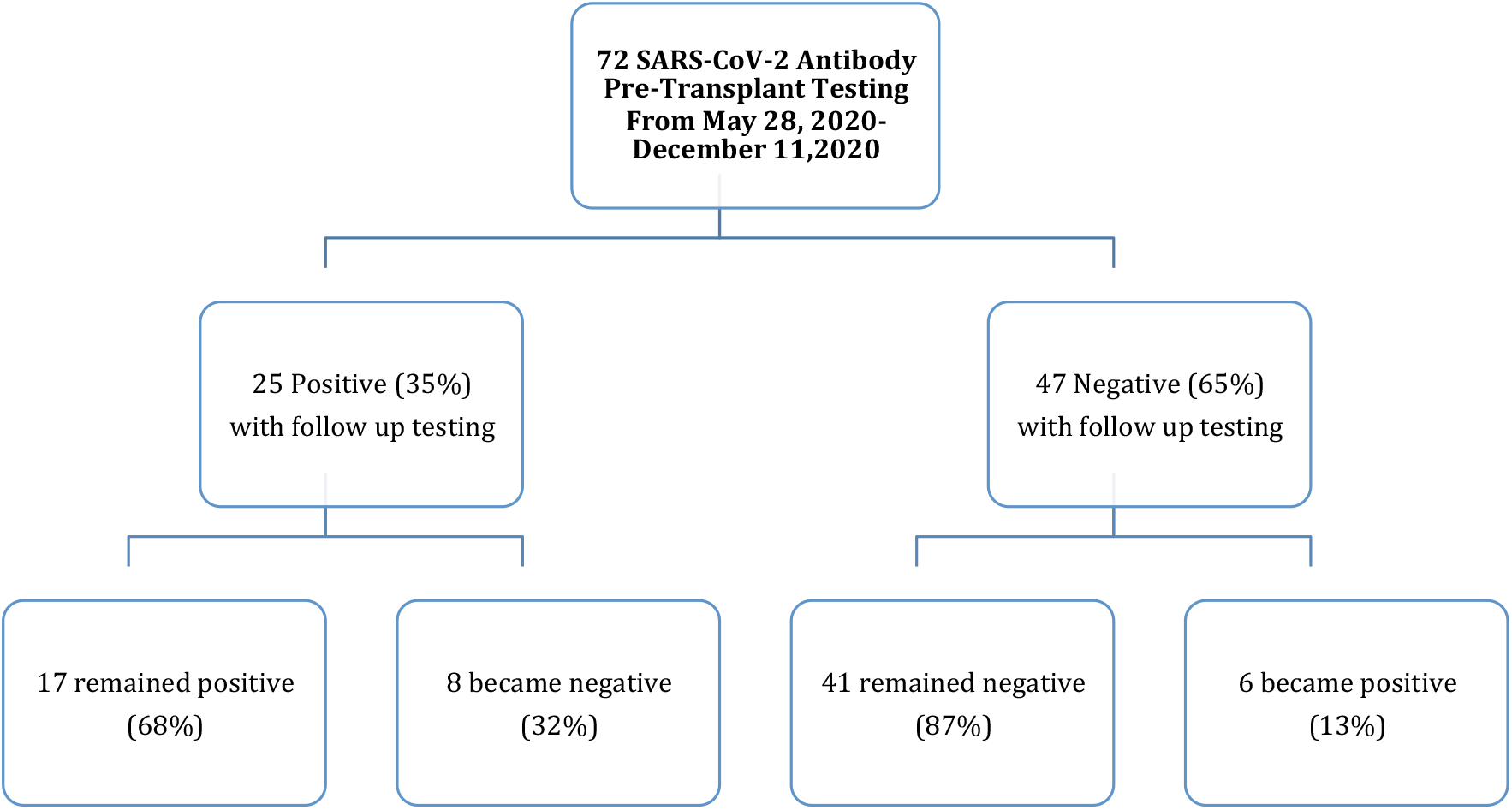
Flow-sheet of Patient Testing for SARS-CoV-2 Antibody. Figure 1 demonstrates the antibody testing results of 72 kidney transplant recipients who received SARS-CoV-2 antibody testing both before and after transplant and follow up testing results.

### Laboratory Evaluation

#### SARS-CoV-2 Antibody Testing

All patients in the study were initially screened for SARS-CoV-2 antibodies upon admission for kidney transplant in the clinical setting using a clinically validated test targeting the RBD of the S1 spike protein (S-RBD) to detect SARS-CoV-2 antibodies which gives either a positive or negative result.

To do further quantitative analysis on a subset of patients, all patients with a positive test were reviewed for availability of left over sera in our tissue-typing laboratory. The Weill Cornell histocompatibility lab performing donor specific antibody testing stores left over sera for transplant candidates and transplant recipients for 6 months. Any patient with a positive pre-transplant SARS-CoV-2 antibody, who had at least 1 pre-transplant and 1 post-transplant stored sample available was included in the quantitative analysis (n=10). In addition, sequential sera samples from 4 wait-listed dialysis patients with SARS-CoV-2 antibodies was also available for testing. A detailed description of these patients is presented in Table 2.

**Table 2:** Characteristics of Patients for Semi-Quantitative Analysis.

| **Characteristics of Patients for Semi-Quantitative Analysis** | **Dialysis**<br>**(n=4)** | **Transplant**<br>**(n=10)** |
| --- | --- | --- |
| **Samples** | **(N=12)** | **(N=26)** |
| **Age, mean (SD)** | **62.5 (3)** | **52.7 (13.6)** |
| **Gender N, (%), Female** | **3 (75)** | **3 (30)** |
| **Race N (%)** |  |  |
| **Caucasian** | **1 (25)** | **1 (10)** |
| **Black** | **3 (75)** | **3 (30)** |
| **Hispanic** | **0 (0)** | **3 (30)** |
| **Asian** | **0 (0)** | **3 (30)** |
| **Induction ? 100% ATG?** |  |  |
| **Symptomatic COVID-19** | **3 (75)** | **7 (70)** |
| **Time from Peak Infections in New York to First Testing (days), mean (SD)** | **155.0 (41.9)** | **108.4 (58.8)** |

Testing for the quantitative 10 transplant recipients and 4 dialysis patients testing was performed using the following assays: 1) measurement of IgG Index value and IgM Index value using the SARS-CoV-2 Pylon 3D analyzer (ET Healthcare) as previously described^6^, 2) SARS-CoV-2 total receptor binding domain (RBD) assay to measure the overall binding between SARS-CoV-2 antibodies and the RBD of the virus S protein, 3) SARS-CoV-2 Antibody Avidity Assay that measures the rate of SARS-CoV-2 specific antibody dissociation from RBD, which is inversely correlated with the antibody avidity, and 4) SARS-CoV-2 Surrogate Neutralizing Antibody Assay (SNAb) which is a competitive binding assay that measures the percentage of RBD-ACE2 binding and inversely correlates with the SNAb binding inhibition (neutralizing activity).

#### Statistical Analysis

GraphPad Prism 9 was used to determine median, mean and standard deviation for all data. Mann-Whitney t-test was used to calculate p values for continuous variables, Wilcoxon t-test was used to calculate p values for paired samples and Fisher’s exact test was used to calculate p values for categorical variables.

## Results

### SARS-CoV-2 Antibody Testing Results of Pre-Transplant Patients

Patient demographics and immunosuppression regimens for all patients, subdivided according to pre-transplant antibody status, is shown in Table 1. Patients in both groups were similar. Follow up creatinine at 1-month, 3-months and 6 months was similar between the two groups (Figure 2). One patient lost their graft in the antibody positive group. Three patients in the antibody negative group developed COVID-19 post transplant while no patients in the antibody positive group had re-infection.

**Figure 2:**
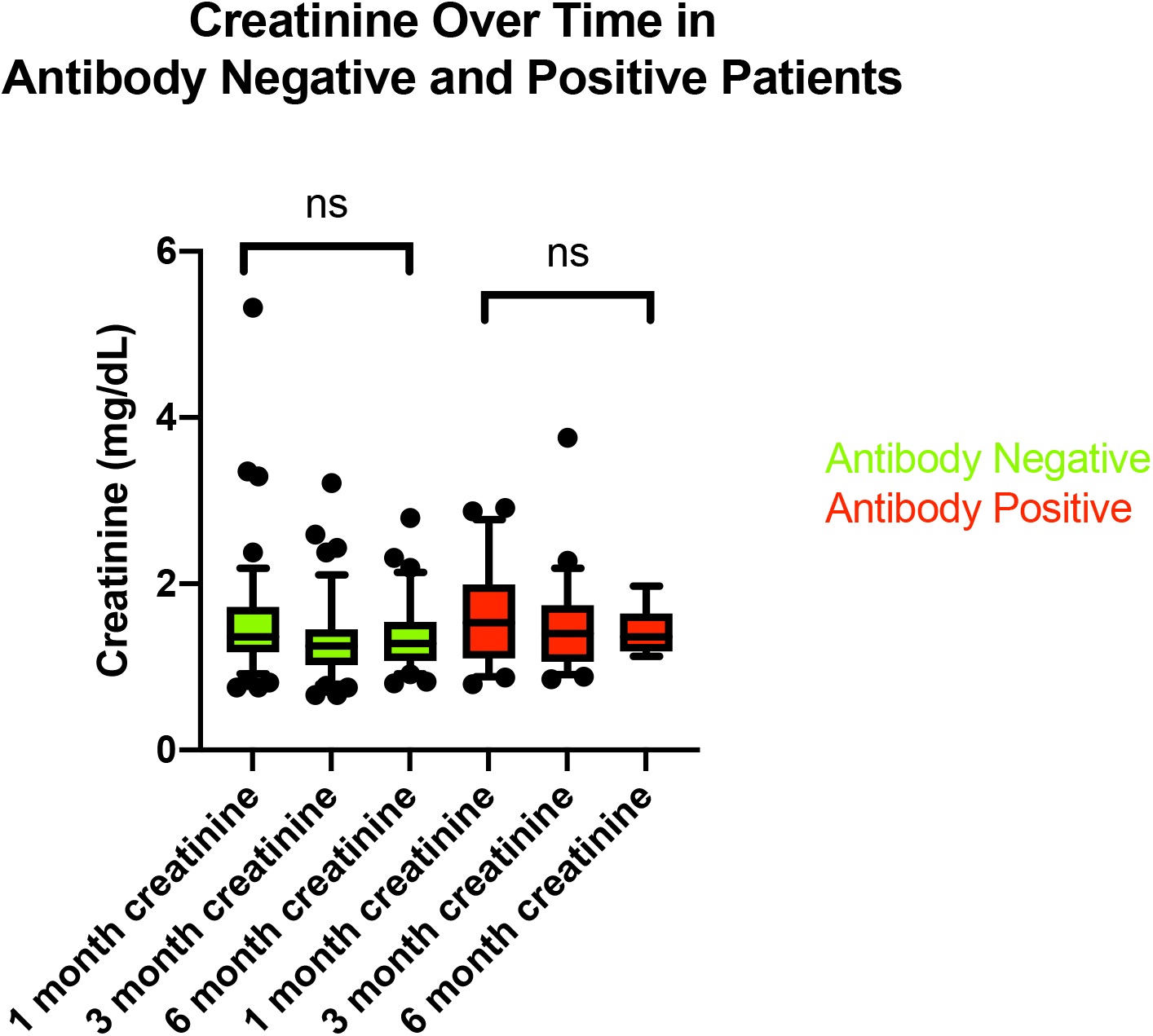
Creatinine Over Time in Antibody Positive and Negative Patients. Box and Whisker plots showing the 10^th^, 25^th^, 50^th^ (median), 75^th^ and 90^th^ percentiles of serum creatinine over time in the patients who had a positive SARS-CoV-2 antibody test pre-transplant and those with a negative SARS-CoV-2 antibody pre-transplant. The change in creatinine over time was not significant in either group.

### Prospective Follow up of Antibody Positive Patients

Of the 25 patients who were antibody positive at the time of transplant, 17 patients (68%) had persistent evidence of antibodies after induction immunosuppression at a median of 83 days post-transplant (range 4, 243). All 8 patients who had a positive test prior to transplant and converted to a negative test post transplant denied a history of COVID-19 like illness and were therefore considered to be asymptomatic for COVID-19. Conversely, of the 17 who had persistently positive tests, 7 (41%) were asymptomatic infections. Patients with symptomatic COVID-19 illness pre-transplant were significantly more likely to have a persistently positive test post-transplant (p=0.01). Figure 3A and 3B demonstrate the change in antibody positivity in patients after transplant and the change in antibody status after transplant in patients with and without symptomatic infection.

**Figure 3:**
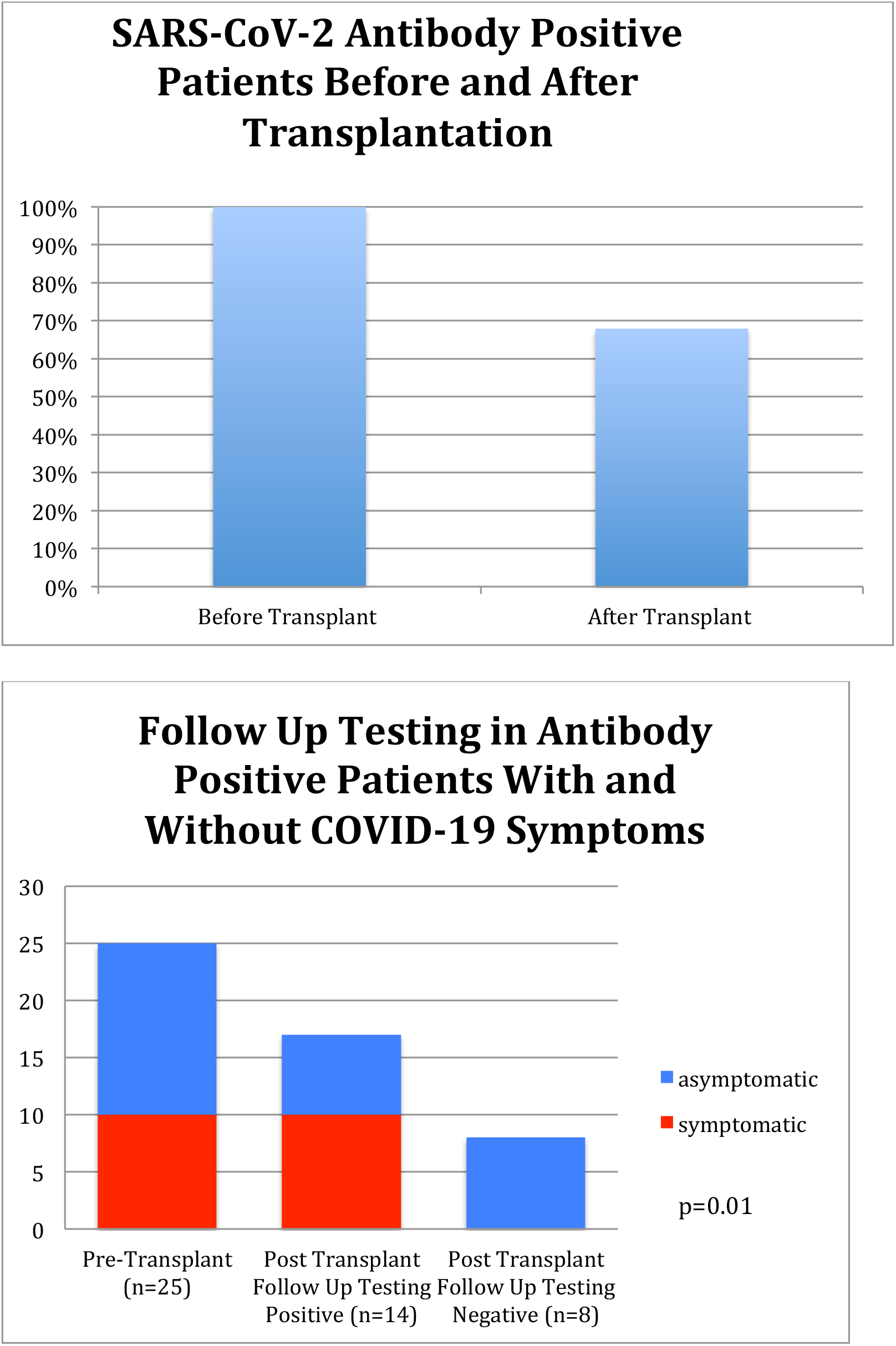
A and B Prospective Follow up of Antibody testing. A demonstrates the number of patients with Positive Antibody Testing at the time of transplant (25) and the percentage of those who remained positive after transplant (17 out of 25, 68%). Figure 3 B demonstrates the percentage of patients with positive antibody tests that remained positive after transplant and whether or not they had a history of symptomatic COVID-19 infection. After transplant, patients were significantly more likely to have a persistently positive test if they had a symptomatic COVID-19 illness (p=0.01).

### Quantitative measurement of antibody titers

Twenty-six samples from 10 patients with history of COVID-19 illness who were transplanted during the study period had semi-quantitative analysis to measure their IgG and IgM levels before and after transplant. SARS-CoV-2 IgG levels were evaluated at 4 time points: 1) pre-transplant, 2) 1-60 days after transplant, 3) 61-120 days after transplant, and 4) more than 120 days after transplant. The percent reduction of IgG from pre-transplant levels at each of the time points was 44%, 72% and 91% respectively and is demonstrated in Figure 4. Box and Whisker Plots showing IgG and IgM levels over time are shown in Figures 5A and 5B.

**Figure 4:**
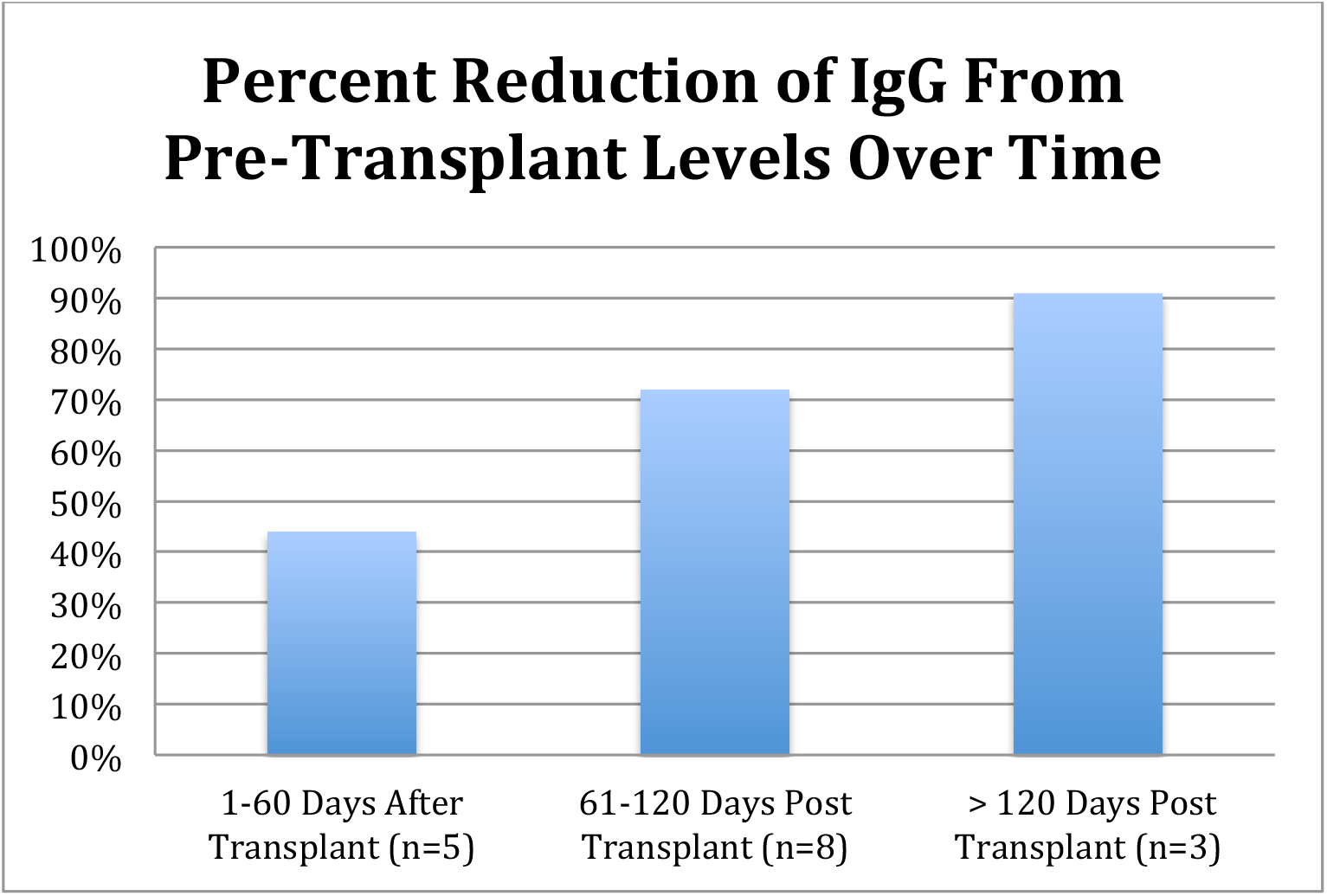
Percentage Reduction of IgG Over Time. Figure 3 demonstrates the percent reduction of median IgG levels from pre-transplant over time. By 4 months there was a 91% reduction in IgG levels, although all patients tested at that time point continued to have a positive Index Value indicating the presence of antibodies to SARS-CoV-2.

**Figure 5A and B.**
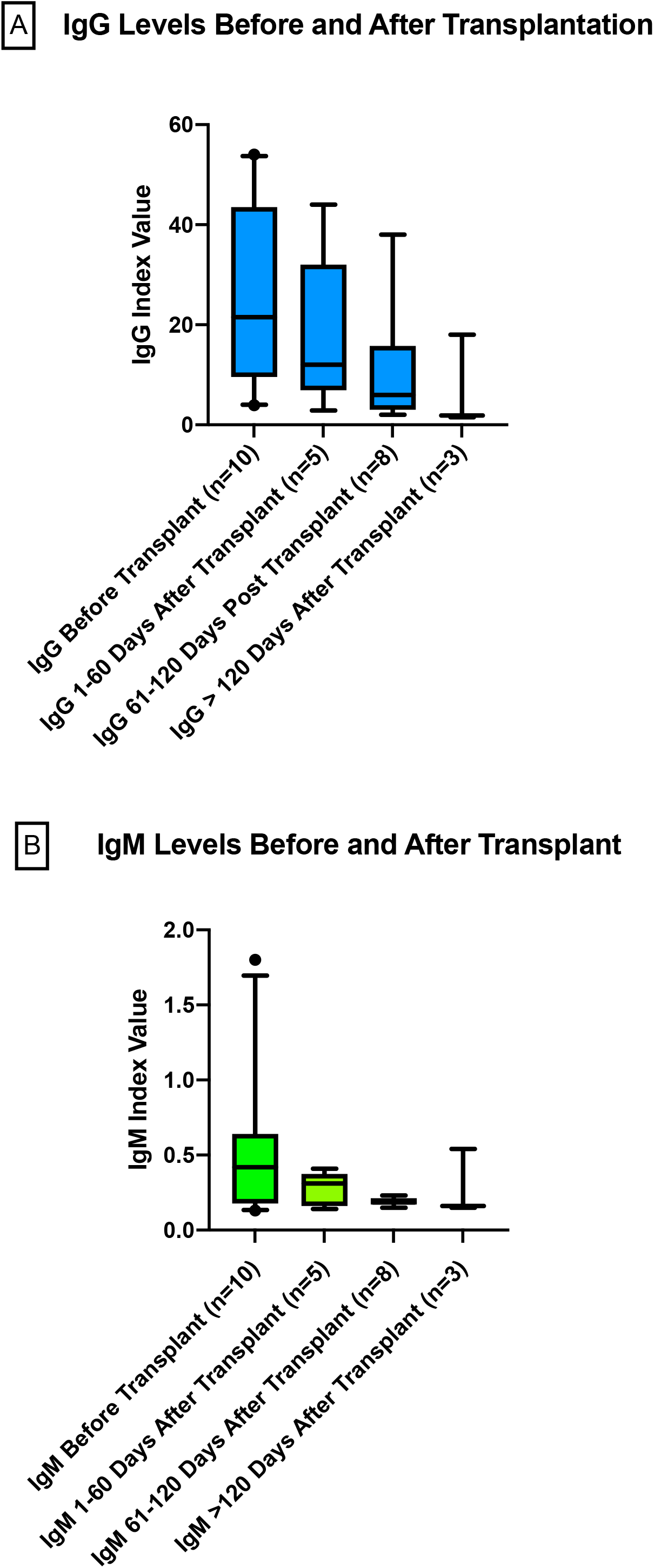
Box and Whisker plots showing the 10^th^, 25^th^, 50^th^ (median), 75^th^ and 90^th^ percentiles of IgG and IgM over time in patients over four time periods: 1) Pre-Transplant 2) 1-60 Days After Transplant 3) 61-120 Days After Transplant and 4) More Than 120 Days After Transplant. A positive value is an Index Value (IV) > 1

### SARS-CoV-2 antibody evaluation in transplant patients compared to wait-listed dialysis patients

To determine if the changes seen in SARS-CoV-2 antibodies over time was similar to that of non-transplant patients, 20 samples from the transplant cohort and stored sera from a cohort of wait-listed dialysis patients with a positive SARS-CoV-2 antibody from the same time frame were tested. Characteristics of patients from both cohorts are listed in Table 2. All transplant patients had a sample taken before transplant and at a mean of 82.4 ±29 days after transplant. Dialysis patients had 2 samples drawn at a mean of 60 ±7 days between 2 samples. The time difference between samples in the transplant cohort and dialysis cohorts were not significantly different (p=0.34). Similarly, initial samples from both the transplant and dialysis cohorts were taken during the same time period after the peak of infections in New York City (mean 108 ±59 after the peak infections in New York City for transplant cohort and for wait-listed dialysis cohort 155 ±7 days, p = 0.17).

Figure 6 demonstrates Box and Whisker Plots comparing antibody analysis in transplant and dialysis patients of the IgG over time (panel A), total receptor binding domain antibody assay demonstrating the binding between SARS-CoV-2 antibodies and the RBD of the virus S protein (panel B), avidity over time (panel C) and the neutralizing antibody over time (panel D). For avidity (panel C) the dissociation measured is inversely correlated to antibody avidity (i.e. the decrease over time shown demonstrates an increase in avidity over time) and for the neutralizing antibody (Panel D), the percentage of RBD-ACE-2 binding is inversely correlated with neutralizing activity and therefore the increase shown over time in the transplant recipients signifies a decrease in neutralizing activity over time. For all assays, there was no significant difference in baseline measurements between pre-transplant values and dialysis cohorts (p=0.54, p=0.95, p =1.0, and p=0.19 respectively). Paired t-tests for all assays comparing before transplant and after transplant measurements demonstrated a significant decline for IgG over time, total receptor binding domain assay over time, and neutralizing antibody over time, while a significant increase in avidity over time was found. The percent reduction of IgG in transplant patients (53%) was significantly greater than the reduction for dialysis patients (14%) (p=0.008). The percent reduction in the total receptor binding domain assay for transplant patients (75% reduction) was significantly greater than the reduction for dialysis patients (1.5%) (p=0.002). There was no significant difference between the percent changes seen in transplant and dialysis patients for the avidity of the antibody (19.5% increase in transplant versus 6% increase in dialysis, P=0.08) or the neutralizing capabilities of the antibody (119% decrease in transplant versus 16.5% decrease for dialysis patients, P=0.43).

**Figure 6:**
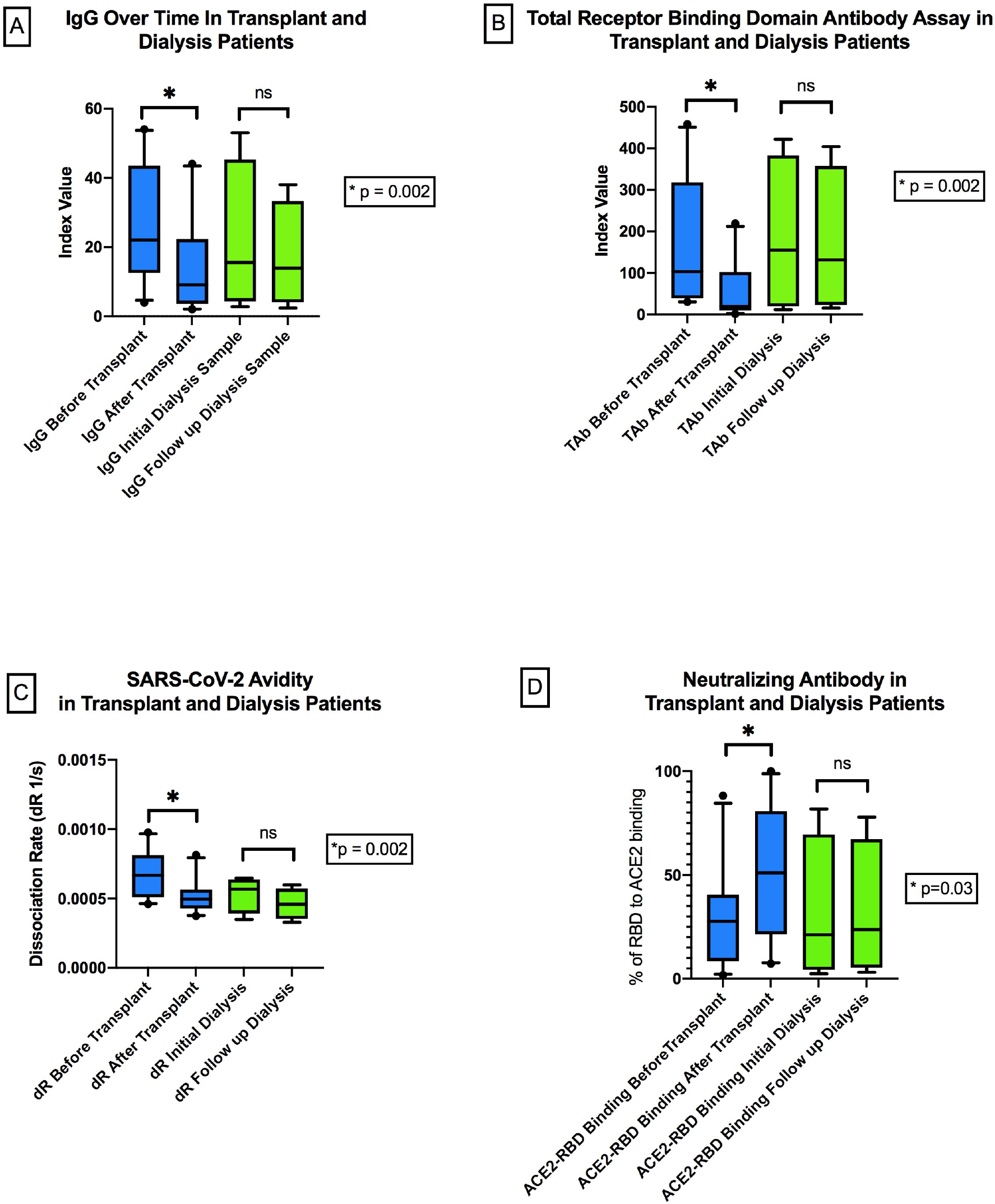
SARS-CoV-2 Antibody Response in Over Time in Transplant and Dialysis Patients. Panel A: Box and Whisker plots showing the 10^th^, 25^th^, 50^th^ (median), 75^th^ and 90^th^ percentiles for the IgG in transplant and dialysis patients over time. A positive value is an Index Value (IV) > 1. Panel B: Box and Whisker plots showing the 10^th^, 25^th^, 50^th^ (median), 75^th^ and 90^th^ percentiles for the Total Receptor Binding Domain (RBD) Antibody Assay (TAb) that measures the overall binding between SARS-CoV-2 antibodies and the RBD of the virus S protein. A positive value is an Index Value (IV) > 1. Panel C: Box and Whisker plots showing the 10^th^, 25^th^, 50^th^ (median), 75^th^ and 90^th^ percentiles of the avidity assay that measures the rate of SARS-CoV-2 specific antibody dissociation from RBD, which is inversely correlated with the antibody avidity. The Y axis represents the relative dissociation rate (dR) which is calculated by fitting the first order rate equation to the dissociation profile: Ln(Signal_t / Signal_0) =Ln([bound]/[total])=-dR t. Panel D: Box and Whisker plots showing the 10^th^, 25^th^, 50^th^ (median), 75^th^ and 90^th^ percentiles of the Neutralizing Antibody (SNAb) that is based on the SARS-CoV-2 antibody-mediated inhibition of the interaction between the ACE2 receptor protein and the RBD. The Y-axis represents the percentage of RBD-ACE2 binding and is measured as %B/B0 = [sample relative fluorescence unit (RFU)/negative blank (RFU)] *100%, which is inversely correlated with antibody neutralizing activity. Mean time from Pre Transplant Sample to Post Transplant Sample was 84 days, while the mean time from Initial Dialysis to Follow up Dialysis Sample was 60 days. The difference in time between the initial and follow up samples in the Transplant and Dialysis Cohorts was not significant (p=0.34).

## Discussion

In this study we evaluated SARS-CoV-2 antibody levels before and after the initiation of induction and maintenance immunosuppression in kidney transplant recipients. This is the first report to date to describe the antibody response in patients who recently received induction immunosuppression for kidney transplant. When compared to wait-listed dialysis patients who had positive testing for SARS-CoV-2 antibody during a similar time period, transplant patients had a significantly greater decline in antibody levels after the initiation of immunosuppression, suggesting that induction therapy at the time of transplantation does impact SARS-CoV-2 antibody titers. Similarly our analysis of the quality of the immune response over time, as measured by the receptor binding domain assay, antibody neutralizing assay, and avidity assays again demonstrated a more pronounced effect over time in patients on immunosuppression compared to those on dialysis.

Although high rates of SARS-CoV-2 infections have been reported in transplant candidates and recipients^7^, little is known about how durable immunity is in the setting of transplant immunosuppression. Our study demonstrates that at a median of 83 days after transplant, 68% of patients still had positive antibodies, indicating a lasting response in most patients. We also demonstrated that patients with a symptomatic infection were significantly more likely to have a durable antibody response after transplant. Our data reaffirms previous reports demonstrating that SARS-CoV-2 antibody levels are stronger in patients with symptomatic infection, even in the transplant population^8^. Additionally, no patients in the group with positive antibody testing pre transplant had a COVID infection post transplant, while 3 patients with a negative antibody developed COVID after transplant.

Despite the short follow up, 32% of patients showed waning immunity, although the more granular analysis did confirm an appropriate maturation and development of immune response. Our data is similar to other reports including the study by Chavarot et al. that measured the IgG levels of 42 kidney transplant recipients at 2 and 6 months after confirmed COVID-19 illness and demonstrated at 2 months 71.4% of patients continued to be IgG positive while at 6 months this number dropped to 36.4%^9^. This is also consistent with a recent a report of solid organ transplant recipients that demonstrated that patients with a higher level of immunosuppression (greater than 2 agents) were less likely to have an antibody response^10^. Although vaccination efforts may curb the risk for COVID-19 illness, data is emerging describing the immune response to COVID vaccination in the transplant population. Boyarsky and colleagues recently published data on efficacy of vaccination in 658 transplant recipients who received 2 doses of one of the approved mRNA vaccines and demonstrated that only 54% had a response after 2 doses of vaccine^11^. Their work demonstrates the need to better understand patient characteristics that predict long-lasting immunity post vaccination and to follow patients closely even after receiving 2 doses of an approved vaccine.

Immunosuppressed patients may not mount an effective immune response to the vaccine and this may be dependent on variety of factors. These factors require careful prospective studies that are ongoing. The optimal timing for vaccination, measuring antibody titers after transplantation, and risks and benefits of a third dose remain to be determined.

Our quantitative analysis of transplant and wait-listed dialysis patients demonstrates that in both cohorts IgG levels decrease over time. To our knowledge this is first report to compare antibody response in dialysis and transplant patients. Our results are also consistent with previous studies in non-transplant populations that demonstrate a decline in IgG antibody levels over time ^12-15^. Yet when comparing the decline in IgG between transplant and dialysis patients, we demonstrated that the percent decrease in kidney transplant recipients was significantly greater over the time period which the transplant recipients received induction immunotherapy and maintenance immunosuppression. We also saw a significant decline in the binding of the antibody to the receptor binding domain for the transplant patients compared to dialysis patients, however, the antibody avidity and neutralizing antibody activity were not significantly different between the two groups (despite showing a significant change in transplant patients over time). Potentially a significant difference was not seen in the neutralizing antibodies and avidity assays due to the small sample size of both cohorts. Overall the general patterns seen in the kidney transplant population and dialysis population mirror what has been shown in previous studies in non-transplant populations^14,16-20^in that both show increased avidity of SARS-CoV-2 antibody over time, which reflects both affinity maturation and multivalent binding development after infection. We also observed increased antibody avidity over time, which is consistent with the notion of continued evolution of the humoral immune response and supported by previous evidence that memory B cell response continues to evolve and express antibodies with increased neutralizing potency and breadth^14,15^. The extent to which immunosuppression plays a role in these particular responses is more difficult to assess given the small sample size, however, it does appear the response in transplant patients overall is attenuated when compared to the dialysis controls. Larger studies are needed in both transplant and dialysis patients to better evaluate these findings.

Our data set has several limitations. First, there were only a small number of samples that were available for sequential testing and there was only a short duration of follow-up. In the cohort of patients who underwent quantitative and qualitative testing, all received lymphocyte depleting induction therapy and therefore it is unknown whether results would be similar if patients had another type of induction therapy, nor could we compare whether the attenuated response would also be seen in patients who underwent induction with IL-2 receptor blocking agents. Such data is important as some have suggested using less potent immunosuppression, especially for induction therapy in the COVID era. Finally, dialysis patients are known to have an impaired immune response^21^ and therefore the study would be strengthened by having a non dialysis cohort to compare our findings as well. It is reassuring, however, that overall our findings mirror that done by other centers in both transplant and non-transplant patients.

Overall, this is the first study to describe the changes in humoral immunity in kidney transplant recipients pre and post-transplantation. We have demonstrated that (1) there is significant heterogeneity in the durability of the humoral response to SARS-CoV-2 in the transplant population and (2) transplant patients experience a significantly greater decrease in IgG levels directed against SARS-CoV-2 RBD protein during the first year as compared to wait-listed dialysis patients studied over a similar time period. Our data suggest that immunity in vaccinated kidney transplant candidates should be followed closely after transplant. Validation of these findings and identifying the basis for the heterogeneous response should be studied in a larger prospective multi-center investigation.

## Data Availability

all data is available upon request

## Acknowledgements

We would like to thank the entire Division of Nephrology and Transplantation for their intellectual contributions to the manuscript, and specifically Dr. Manikkam Suthanthiran for his guidance and mentorship. In addition we would like to thank the Immunogenetics and Transplantation Laboratory at Weill Cornell for storing and allowing use of the samples for analysis in this project and Dr. Vijay Sharma for his contribution to this work.

## Author Contributions

ML, ZZ, MC and DD all contributed to the study design, data collection, data analysis, writing and editing of the manuscript. ZZ, AS and MC performed the antibody testing for quantitative and qualitative analysis. SR assisted in data collection and analysis. SS, ZK, SA, NH, JM-P, RC-S, JRL, TS, MA, SK all contributed to data collection and editing of the manuscript.

